# Antibiotic susceptibility patterns of pathogens isolated from laboratory specimens at Livingstone Central Hospital in Zambia

**DOI:** 10.1101/2022.05.19.22275336

**Authors:** Thresa N. Mwansa, Kingsley Kamvuma, John Amos Mulemena, Christopher Newton Phiri, Warren Chanda

**Affiliations:** Mulungushi University School of Medicine and Health Sciences, Department of Pathology and Microbiology, Livingstone Zambia

**Author notes:** **Corresponding author:** Mr. Warren Chanda, Mulungushi University School of Medicine and Health Sciences, Department of Pathology and Microbiology, Livingstone Zambia.

**Keywords:** Antibiotic resistance, Livingstone Central Hospital, Imipenem, *Staphylococcus aureus*, Ampicillin

## Abstract

**Background:** Antibiotics are essential commodities in managing bacterial infections in humans, animals and plants but are hampered by the development of antibiotic resistance which is one of the most serious public health threats of the twenty-first century. Moreover, the rate at which novel antibiotics are discovered is slower that the rate of emerging antibiotic resistance. Therefore, the few remaining potent antibiotics in clinical setting should be safeguarded by closer monitoring of their effectiveness via periodic antibiogram studies. This study aimed to evaluate the antibiotic susceptibility patterns of routinely isolated bacteria at Livingstone Central Hospital (LCH).

**Methods:** A cohort retrospective study with secondary information collected from electronic laboratory system generated reports on all isolated organisms at LCH microbiology laboratory for three years (January 2019 to December 2021) was used. Study variables such as age, gender, patient’s location, name of the organism and the antibiotic susceptibility were considered. Descriptive statistics was used to describe our data and a chi-square test was used for categorical variables where a p-value of ≤0.05 was considered as statistically significant.

**Results:** A total of 765 specimens were processed from January 2019 to December 2021 and only 500 (65.4%) met the inclusion criteria for this study. Of the 500, 291(58.2%) specimens were received from female and from the age-group 17-39 years (253, 50.6%) and 40-80 years (145, 29%) in form of blood (331, 66.2%), urine (165, 33%) and sputum (4, 0.8%). The out-patient department (323, 64.6%) had a higher number of specimen culture requests that reduced from 175 (35%) for the year 2019 and 2020 to 150 (30%) for year 2021. Amongst the common bacterial isolates identified, *Staphylococcus aureus* (142, 28.4%) was the commonest isolate followed by *Escherichia coli* (91, 18.2%), *Enterobacter agglomerans* (76, 15.2%), and *Klebsiella pneumoniae* (43, 8.6%). The resistance pattern indicated that ampicillin (93%) was the least effective drug followed by oxacillin (88%), penicillin (85.6%), co-trimoxazole (81.5%), erythromycin (71.9%), nalidixic acid (68%), ceftazidime (60%), tetracycline (55.1%), and ciprofloxacin (45.9%) whereas the most effective antibiotics were imipenem (14.5%), piperacillin/tazobactam (16.7%) and clindamycin (34.5%). The resistance levels were affected by patient gender, location, and specimen type.

However, the screening of methicillin resistant *Staphylococcus aureus* (MRSA) with cefoxitin showed 76.3% (29/38) susceptibility and 23.7% (9/38) resistance.

**Conclusion:** The commonest bacterial isolates were *Staphylococcus aureus, Escherichia coli, Enterobacter agglomerans, Klebsiella pneumoniae* and *Klebsiella oxytoca*. The least effective antibiotics were ampicillin, penicillin, oxacillin, cotrimoxazole, and erythromycin whereas the most effective antibiotics were imipenem, piperacillin/tazobactam, and clindamycin. Therefore, re-establishing of the empiric therapy is needed for proper patient management, studies to determine the levels of extended spectrum beta lactamase- and carbapenemase-producing bacteria are warranted.

## Introduction

Antibiotics are essential commodities in managing bacterial infections in humans and animals. However, the spread of antibiotic-resistant bacteria had been increased by the overuse and misuse of antibiotics as well as social and economic factors (1). Therefore, antibiotic resistance is a public health concern that has been recognized globally. Studies have indicated that a lot of lives are lost due to antimicrobial resistance related illnesses every year, and almost 100% β-lactams and some 3rd generation cephalosporins and carbapenems have recorded resistance by some organisms (2). Prior to the development of the first β-lactam (penicillin) and its release in medical practice, emergence of resistance had been reported and the first β-lactamase enzyme was identified in *Escherichia coli* (3). Extended Spectrum β-lactamases (ESBLs) are enzymes that deactivate a variety of β-lactam antibiotics including penicillins, cephalosporins (3rd and 4th generations) and monobactams but less likely to deactivate cephamycins like cefoxitin (3-5). Furthermore, the production of carbapenemase enzymes by some Enterobacteriaceae is alarming as these enzymes can break down antibiotics including carbapenem antibiotics, which are typically reserved to treat multidrug-resistant bacterial infections (6). Carbapenem-resistant Enterobacteriaceae (CRE) causes very hard to treat infections because of being resistant to carbapenems that are used on multidrug resistant strains (7). It is sufficed to note that the more bacteria get exposed to antimicrobial agents the more resistance develops. Therefore, there is need to preserve the current antimicrobial agents by using them judiciously otherwise, we risk having pandemic AMR infections that will devastate the global community.

In Africa, especially in low and medium-income countries (LMIC), antimicrobial resistance monitoring is inadequate, but the extensive usage of antibiotics to prevent and treat infectious diseases has led to the emergence and spread of antibiotic resistance which has influenced a particular force on susceptible bacteria leading to resistant strain survival, consequently increasing medical costs, illnesses and deaths of patients (8). Therefore, antibiogram studies in LMIC are important to closely monitor the trends of AMR at hospital and national levels. This retrospective study aimed to assess antibiotics resistance among commonly isolated bacteria from routine specimens at Livingstone Central Hospital from 2019 to 2021.

## Material and Methods

### Study design and site

A retrospective cohort study was conducted at Livingstone Central Hospital (LCH) on routine specimen isolates from paediatric and adults who visited the hospital between January 2019 – December 2021. Livingstone Central Hospital has different departments that include laboratory, internal medicine, surgery, paediatrics, and obstetrics and gynaecology. The LCH microbiology laboratory participates in a bacteriology External Quality Assessment (EQA) program and has been accredited by the Southern African Development Community Accreditation Service (SADCAS).

### Eligibility criteria

This study included all specimens having information on the patient’s gender, age, location (ward/clinic), name of organism and antibiotic susceptibility testing. However, any isolated organism without a species name, and with unknown source (i.e., lack of age-, location-, and gender of patient, and sample type) were excluded from the study.

### Data collection and analysis

Data from electronic laboratory system generated reports on all isolated organisms at LCH microbiology laboratory for three years (January 2019 to December 2021) was used from which information such as age, gender, patient’s location, name of the organism and the antibiotic susceptibility were considered. The collected data was entered, assorted, and coded using Microsoft Excel 2019 and then exported to Statistical Package for Social Science (SPSS) version 20 for analysis. Descriptive statistics was used to describe our data. Graph prism 5 was used for graph generation. A chi-square test was used for categorical variables and a p-value of ≤0.05 was considered statistically significant.

### Ethical consideration

This is a retrospective study that analysed secondary data generated from routine laboratory specimens from both adult and paediatric departments. There was no human interaction, and no personal identifiers were included in this study. However, ethical waiver was granted by Mulungushi University School of Medicine and Health Sciences Research Ethics Committee (SMHS-MU2-2021-33v1) while permission to use the Disa*Lab system generated data was obtained from Livingstone Central Hospital management.

## Results

### General characterization of isolated bacterial organisms

A total of 765 specimens were processed from January 2019 to December 2021 and only 500 (65.4%) met the inclusion criteria for this study. Of the 500, 291(58.2%) specimens were received from female and 209 (41.8%) from males, ranging age between 0 and 80 years with the mean average of 32.3±20.6 years. Categorizing the age groups in 0-16 years, 17-39 years, and 40-80 years, 102 (20.4%)-, 253 (50.6%)-, and 145 (29%)-specimens were received, respectively (Fig. 1A). For the type of specimens requested, blood specimens were 331 (66.2%) followed by urine specimen (165, 33%) and sputum (4, 0.8%; Fig. 1A). Most specimens came from out-patient department (323, 64.6%) while 177 (35.4%) came from the in-patient department (IPD), and 175 (35%) specimens were analysed in the year 2019 and 2020, while 150 (30%) specimens were processed in year 2021 (Fig. 1A). Amongst the common bacterial isolates identified, *Staphylococcus aureus* (142, 28.4%) was the commonest isolate followed by *Escherichia coli* (91, 18.2%), *Enterobacter agglomerans* (76, 15.2%), *Klebsiella pneumoniae* (43, 8.6%), and the least isolated bacteria were *Hafnia alvei, Proteus vulgaris*, and *Yersinia enterocolytica* (3, 0.6% apiece; Fig. 2).

**Figure 1:**
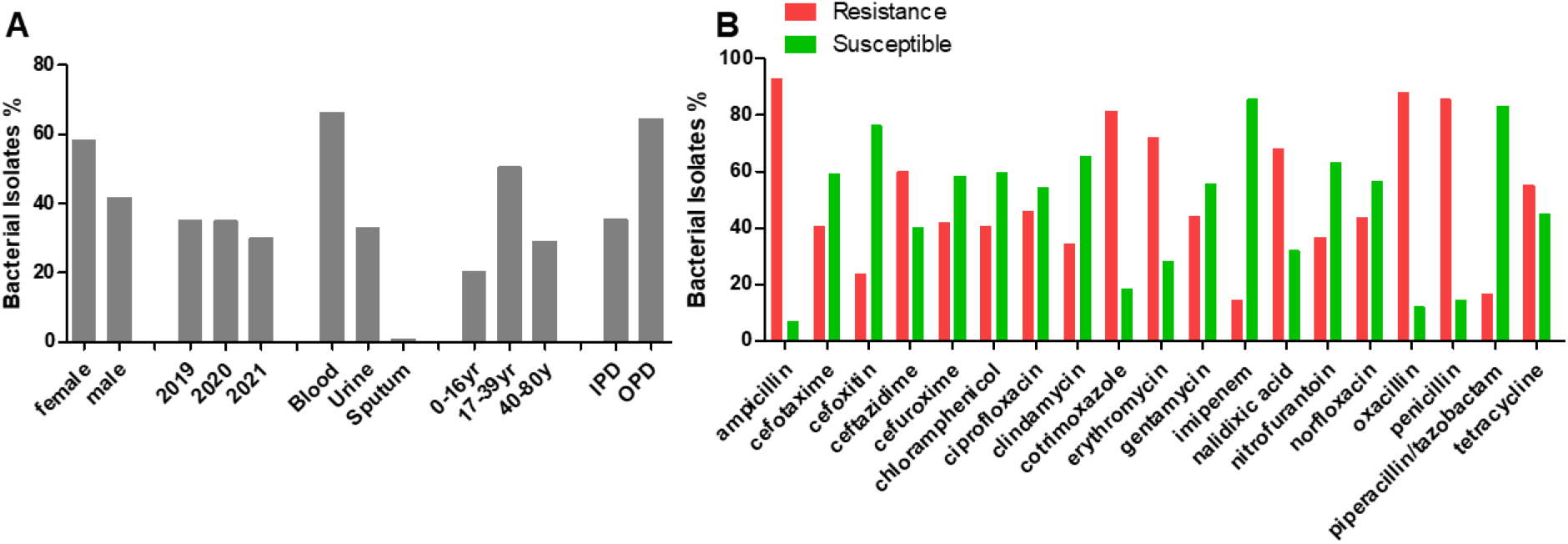
Characterization of bacterial isolates from blood, urine, and sputum specimens. (A) percentage frequencies of isolates based on patient gender, year of isolated, sample type age and location; (B) percentage frequencies of the susceptibility patterns against common utilized antibiotics. IPD: in-patient department, OPD: out-patient department, yr/y: year.

**Figure 2:**
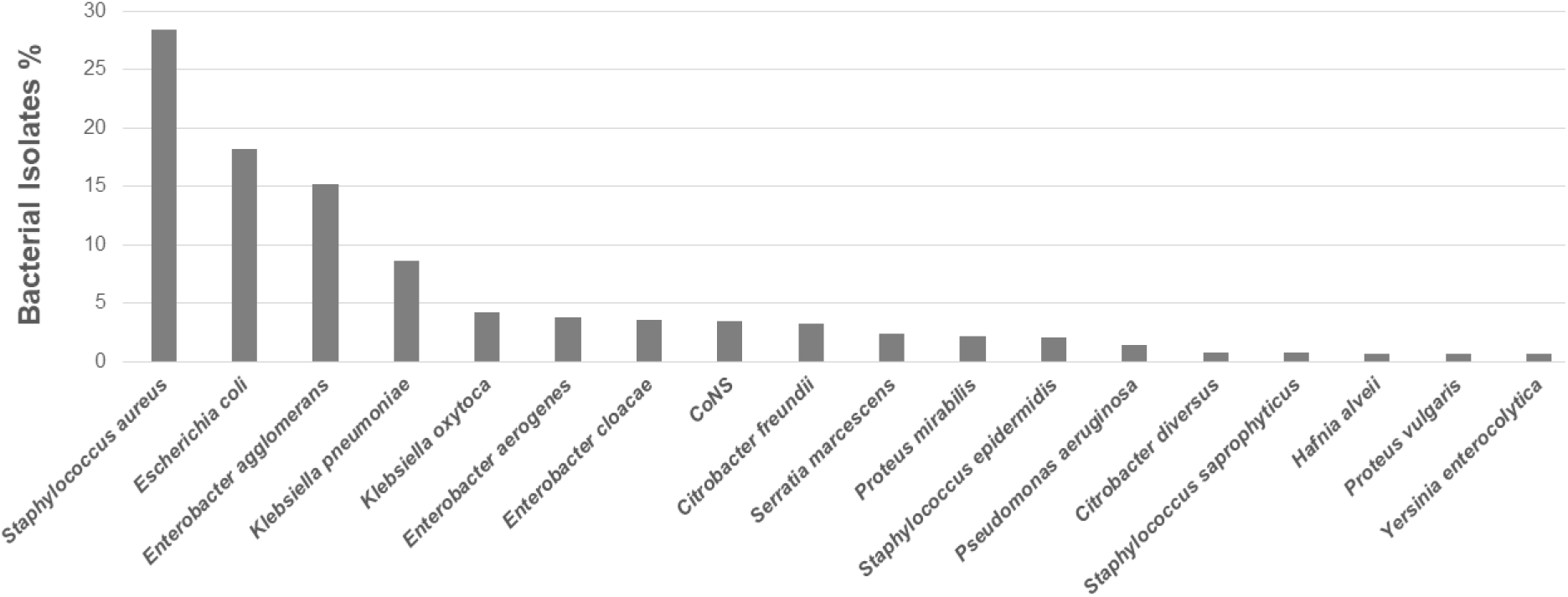
Percentage frequency of bacterial isolate from blood, urine, and sputum specimens. CoNS: Coagulase negative *Staphylococci*.

A large panel of antibiotics that are commonly used at Livingstone Central Hospital (LCH) were used for antimicrobial susceptibility testing following the Clinical and Laboratory Standards Institute (CLSI) recommendations (9). From the list of antibiotics commonly used at LCH, the resistance pattern indicated that ampicillin (93%) was the least effective drug followed by oxacillin (88%), penicillin (85.6%), co-trimoxazole (81.5%), erythromycin (71.9%), nalidixic acid (68%), ceftazidime (60%), tetracycline (55.1%), and ciprofloxacin (45.9%) whereas the most effective antibiotics were imipenem (14.5%), piperacillin/tazobactam (16.7%) and clindamycin (34.5%) as shown in figure 1B. However, the screening of methicillin resistant *Staphylococcus aureus* (MRSA) with cefoxitin showed 76.3% (29/38) susceptibility and 23.7% (9/38) resistance (Fig. 1B).

### The association of patient gender, age group, location, and year of isolation with resistant pattern of bacteria isolates

We conducted a chi-square test to determine the link between resistance pattern of bacteria and independent variables. We found that the potency of both nitrofurantoin and cefuroxime were affected by gender. Specifically, the effectiveness of nitrofurantoin on *Enterobacter agglomerans* (p=0.041), co-trimoxazole on coagulase negative *Staphylococci* (CoNS, p= 0.044), cefuroxime on *Escherichia coli* (p=0.011) and cefotaxime on *Klebsiellla oxytoca* (p=0.026) as presented in Table 1. The patient location impacted negatively on the performance of chloramphenicol and ciprofloxacin against *Enterobacter agglomerans* (p=0.002) and *Klebsiella pneumoniae* (p=0.021), respectively (Table 1). Further analysis revealed that the *Enterobacter aerogenes* resistance to co-trimoxazole (p=0.005) varied with year of isolation, and so was *Staphylococcus aureus* resistance to tetracycline (p=0.009), chloramphenicol (p=0.002) and co-trimoxazole (p=0.041). Lastly, *Staphylococcus aureus* resistance to tetracycline was affected by specimen type (p=0.047; Table 1).

**Table 1:**
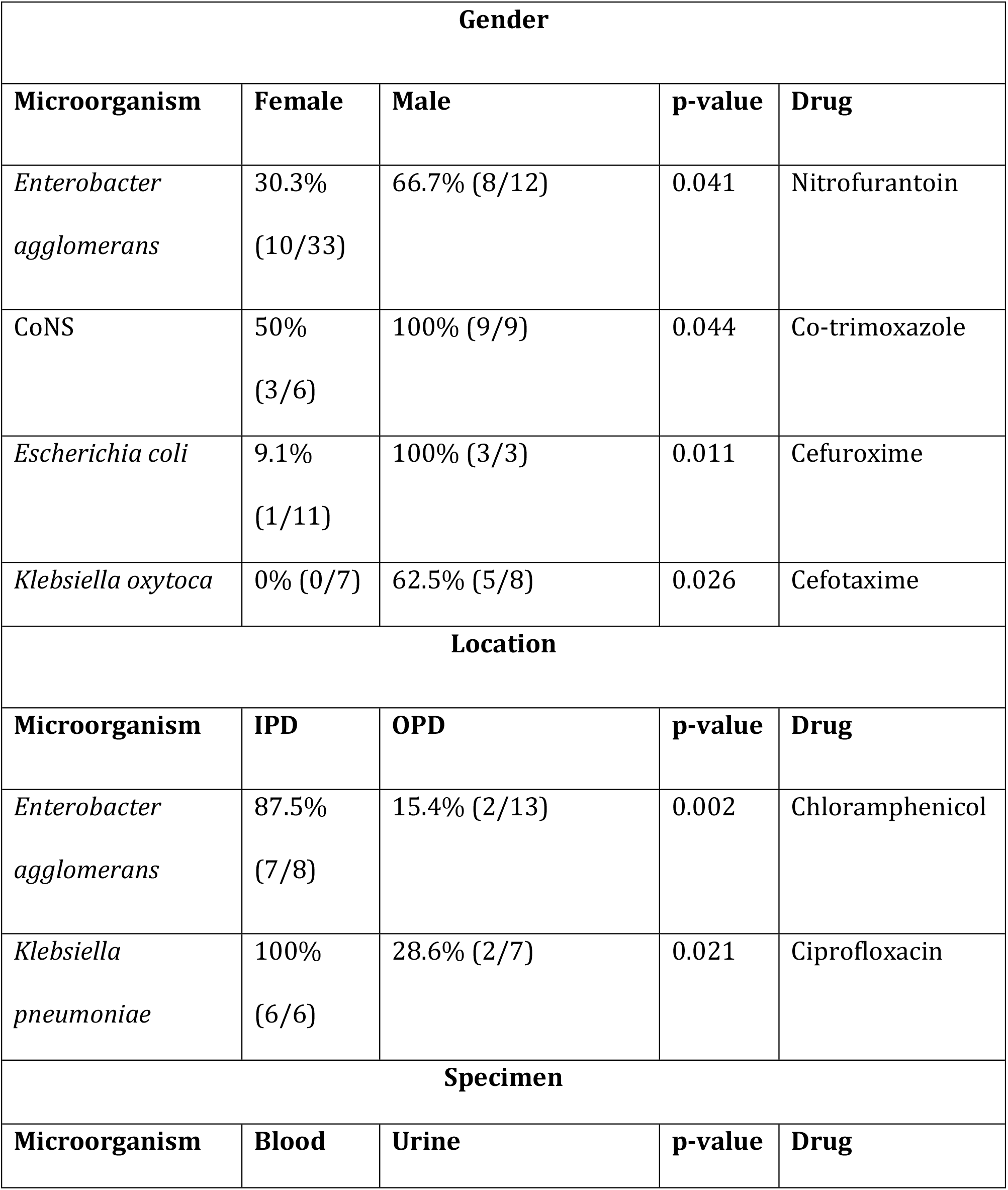
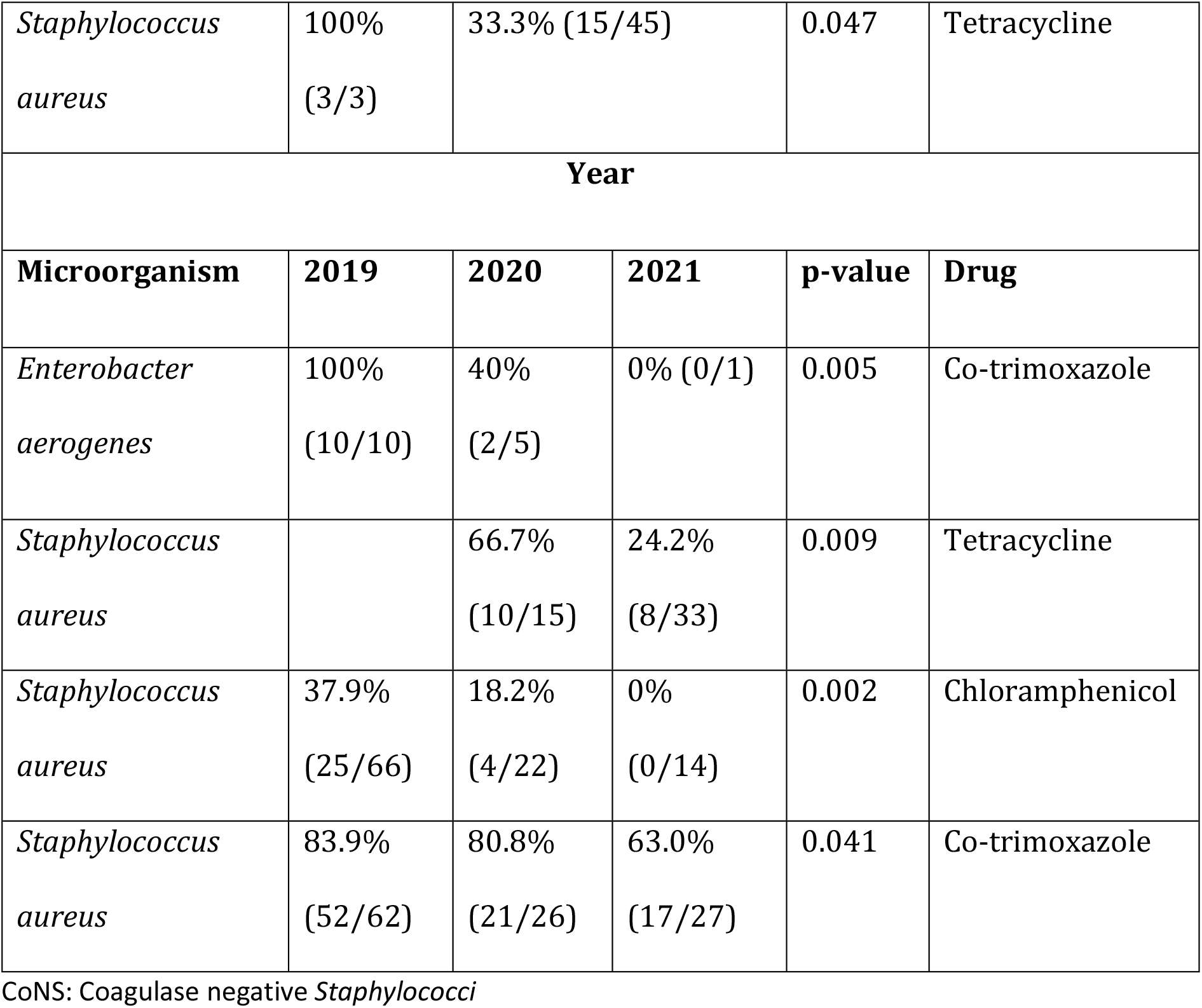
The resistant pattern of some bacteria isolates with respect to patient location, year of isolation, patient gender, and specimen type.

| Gender |  |  |  |  |
| --- | --- | --- | --- | --- |
| Microorganism | Female | Male | p-value | Drug |
| *Enterobacter agglomerans* | 30.3%<br>(10/33) | 66.7% (8/12) | 0.041 | Nitrofurantoin |
| CoNS | 50%<br>(3/6) | 100% (9/9) | 0.044 | Co-trimoxazole |
| *Escherichia coli* | 9.1%<br>(1/11) | 100% (3/3) | 0.011 | Cefuroxime |
| *Klebsiella oxytoca* | 0% (0/7) | 62.5% (5/8) | 0.026 | Cefotaxime |
| Location |  |  |  |  |
| Microorganism | IPD | OPD | p-value | Drug |
| *Enterobacter agglomerans* | 87.5%<br>(7/8) | 15.4% (2/13) | 0.002 | Chloramphenicol |
| *Klebsiella pneumoniae* | 100%<br>(6/6) | 28.6% (2/7) | 0.021 | Ciprofloxacin |
| Specimen |  |  |  |  |
| Microorganism | Blood | Urine | p-value | Drug |

|  |  |  |  |  |  |
| --- | --- | --- | --- | --- | --- |
| *Staphylococcus aureus* | 100%<br>(3/3) | 33.3% (15/45) |  | 0.047 | Tetracycline |
| Year |  |  |  |  |  |
| Microorganism | 2019 | 2020 | 2021 | p-value | Drug |
| *Enterobacter aerogenes* | 100%<br>(10/10) | 40%<br>(2/5) | 0% (0/1) | 0.005 | Co-trimoxazole |
| *Staphylococcus aureus* |  | 66.7%<br>(10/15) | 24.2%<br>(8/33) | 0.009 | Tetracycline |
| *Staphylococcus aureus* | 37.9%<br>(25/66) | 18.2%<br>(4/22) | 0%<br>(0/14) | 0.002 | Chloramphenicol |
| *Staphylococcus aureus* | 83.9%<br>(52/62) | 80.8%<br>(21/26) | 63.0%<br>(17/27) | 0.041 | Co-trimoxazole |

CoNS: Coagulase negative *Staphylococci*

### Characterization of bacterial isolates from paediatric specimens

Out of the total of 500 specimens that met the inclusion criteria for this study, 102 (20.4%) were paediatric specimens. Of the 102, 46(45.1%) specimens were received from female and 56 (54.9%) from males, ranging age between 0 and 16 years with the mean average of 4.12±5.0 years. Many specimens came from in-patient department (76, 74.5%) while 26 (25.5%) came from the out-patient department (OPD), and 37 (36.3%) specimens were analysed in the year 2019, followed by the year 2020 with 29 (28.4%) while 36 (35.3%) were processed in the year 2021 (Fig. 3A). The most isolated bacterium was *Staphylococcus aureus* (44, 43.1%) followed by *Escherichia coli* (17, 16.7%), *Enterobacter agglomerans* (16, 15.7%), *Klebsiella pneumoniae* (7, 6.9%), and *Klebsiella oxytoca* (4, 3.9%; Fig. 3B). Antibiotic susceptibility testing was conducted with a panel of antibiotics that are commonly used at LCH. The resistance pattern revealed that ampicillin (95.5%) was the least effective drug followed by oxacillin (87.5%), penicillin (84.8%), co-trimoxazole (82.5%), erythromycin (74.2%), and nalidixic acid (60%) whereas the most effective antibiotics were imipenem (7.7%), nitrofurantoin (29.6%) and cefotaxime (34.1%) as shown in figure 3C. However, patient gender, patient location, specimen type and year of specimen processing had no statistical effect on the resistance pattern of bacterial isolates.

**Figure 3.**
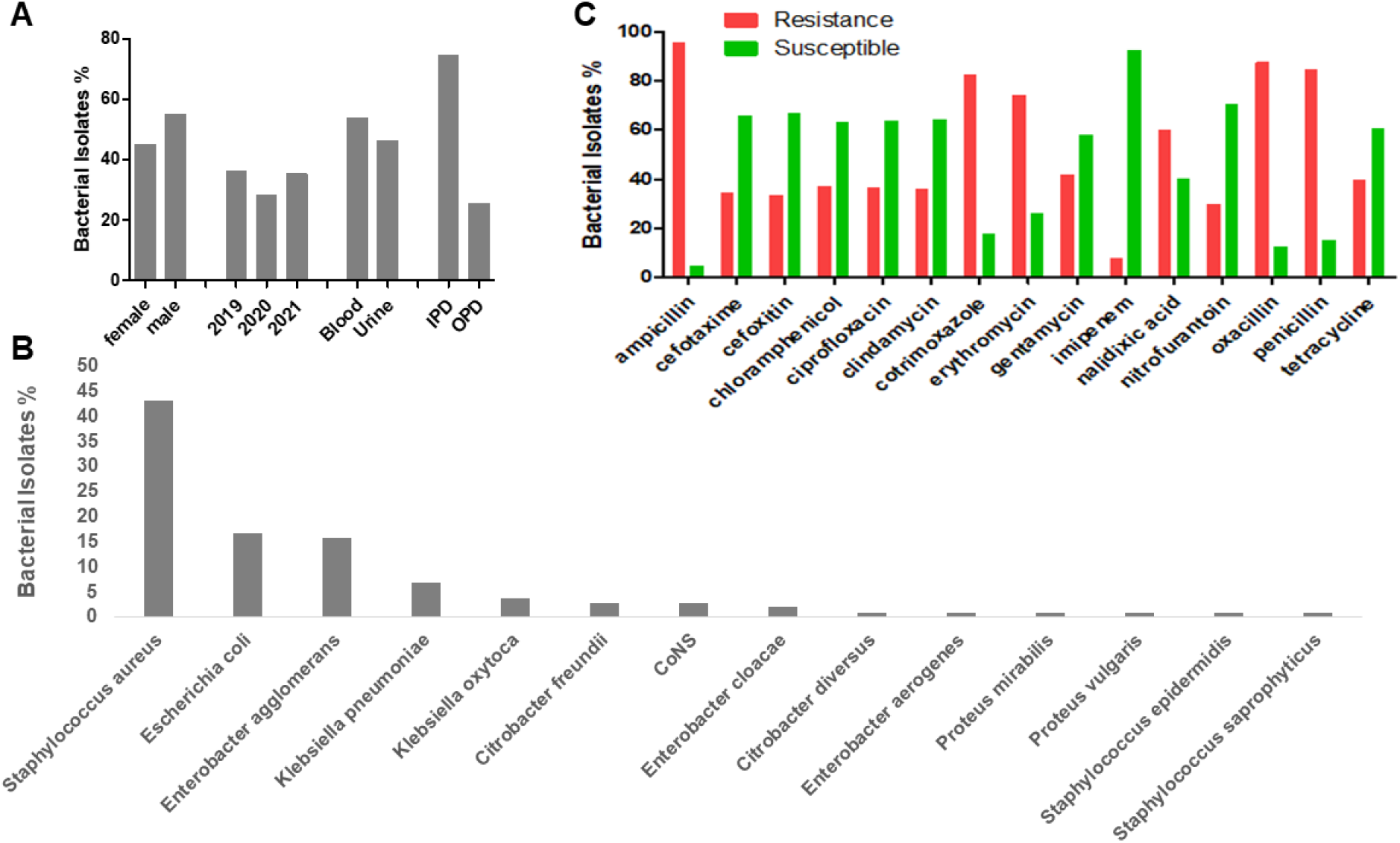
Characterization of bacterial isolates from paediatric specimens; (A) percentage isolates based on patient gender, location, year of isolates, and specimen type, (B) Percentage frequency of bacterial isolate from blood, urine, and sputum specimens, and (C) percentage frequencies of the susceptibility patterns of bacterial isolates. IPD: in-patient department, OPD: out-patient department, CoNS: Coagulase negative *Staphylococci*.

### Characterization of bacterial isolates from in-patient department

Due to the over-use of various disinfectants and drugs in hospitals, bacteria species tend to be highly resistant. So, we wanted to understand the behaviour of bacterial isolates to commonly utilized antibiotics from in-patient department. Out of the 500 specimens that met the inclusion criteria for this study, 177 (34.5%) were specimens from in-patient departments. Of the 177, 92 (52%) specimens were received from female and 85 (48%) from males, ranging age between 0 and 80 years with the mean average of 25.5±24 years. Categorizing the age groups in 0-16 years, 17-39 years, and 40-80 years, 76 (42.9%)-, 58 (32.8%)-, and 43 (24.3%)-specimens were received, respectively (Fig. 4A). For the type of specimens requested, blood specimens were the highest with 113 (63.8%) followed by urine specimen (62, 35%) and sputum (2, 1.2%; Fig. 4A). Also, 62 (35%) specimens were analysed in the year 2021, followed 60 (33.69%)specimens in the year 2020, and 55 (31.1%) specimens in the year 2019 (Fig. 4A). The most isolated bacterium was *Staphylococcus aureus* (50, 28.2%) followed by *Escherichia coli* (32, 18.1%), *Enterobacter agglomerans* (26, 14.7%), *Klebsiella pneumoniae* (12, 6.8%), and *Coagulase Negative Staphylococci* (8, 4.5%) as presented on figure 4B.

**Figure 4.**
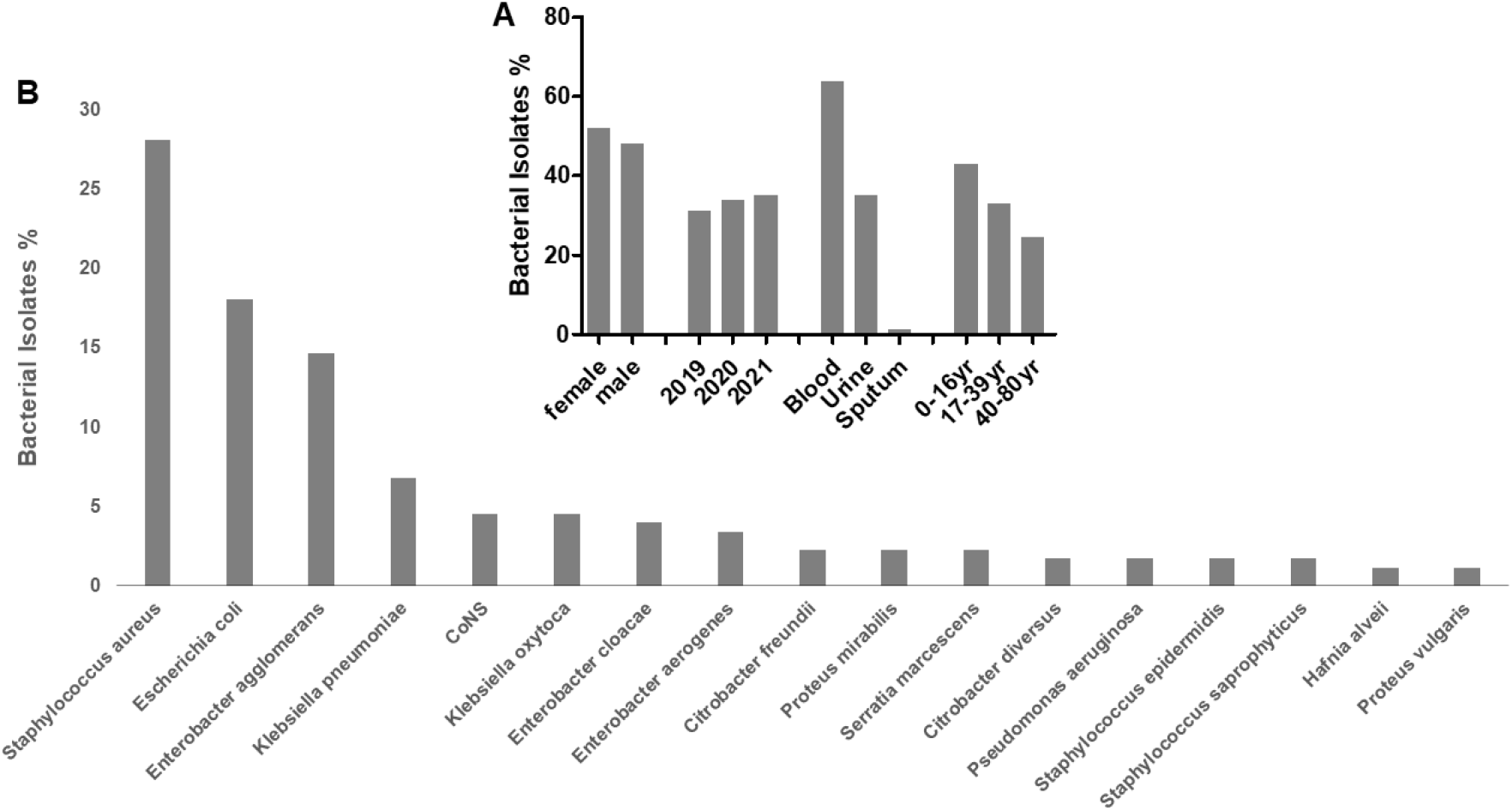
Characterization of bacterial isolates from in-patient department specimens. (A) percentage isolates based on patient gender, year of isolates, specimen type, age category, and (B) percentage frequencies of the susceptibility patterns of isolates from in-patient departments. yr: year.

Antibiotic susceptibility testing was conducted with a panel of antibiotics that are commonly used at LCH. The resistance pattern revealed that ampicillin (93.4%) was the least effective drug followed by penicillin (83%), co-trimoxazole (82.5%), oxacillin (80%), erythromycin (74.4%), nalidixic acid (70.7%), cefuroxime (60%), and tetracycline (48.5%) whereas the most effective antibiotics were norfloxacin (18.2%), imipenem (18.5%) and clindamycin (28.6%) as shown in figure 5. The prevalence of MRSA among *Staphylococcus aureus* that were tested against cefoxitin was 15.4% (2/13) whereas 84.6% (11/13) were methicillin susceptible *Staphylococcus aureus* (MSSA, Fig. 5).

**Figure 5.**
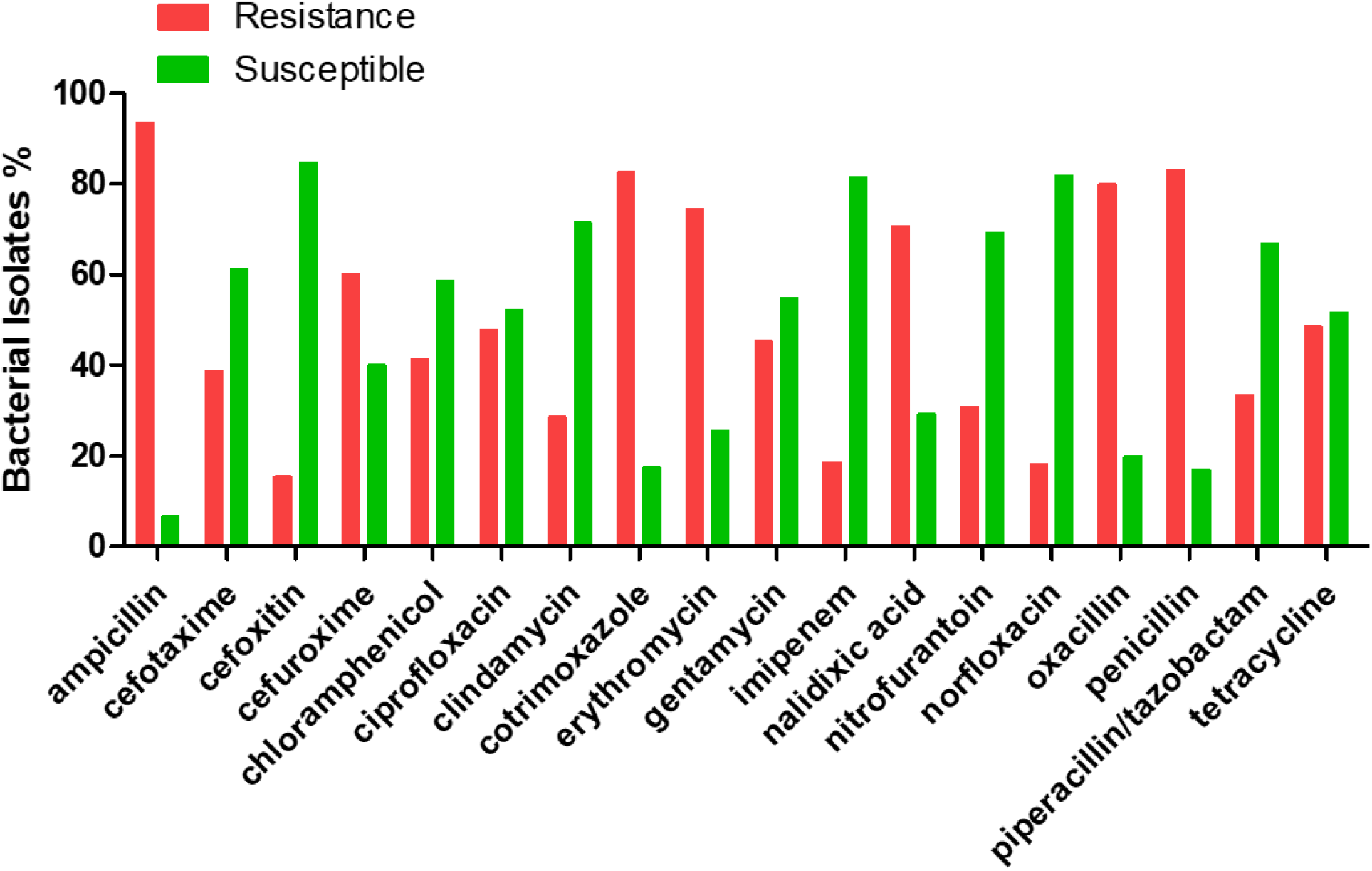
percentage frequencies of the susceptibility pattern of isolates from in-patient departments.

However, *Staphylococcus aureus* resistance to chloramphenicol had a significant reduction from 2019 to 2021 (p=0.006) whereas its resistance to penicillin was the highest in 0-16years age group (p=0.012, Table 2).

**Table 2:** The resistant pattern of some IPD bacteria isolates with respect to year of isolation and patient age group.

| Year |  |  |  |  |  |
| --- | --- | --- | --- | --- | --- |
| Microorganism | 2019 | 2020 | 2021 | p-value | Drug |
| *Staphylococcus aureus* | 52.9%<br>(9/17) | 22.2%<br>(2/9) | 0%<br>(0/11) | 0.006 | Chloramphenicol |
| Age group |  |  |  |  |  |

| Microorganism | 0-16yr | 17-39yr | 40-80yr | p-value | Drug |
| --- | --- | --- | --- | --- | --- |
| *Staphylococcus aureus* | 85.5%<br>(17/19) | 100%<br>(10/10) | 42.9%<br>(3/7) | 0.012 | Penicillin |

yr: year

## Discussion

### General characterization of bacterial isolates from routine specimens

Antimicrobial resistance (AMR) is a global public health problem that requires concerted effort from all sectors of life to control it. Due to lack of data on prevalence rates because of limited or absent antimicrobial surveillance systems and poor resources for antimicrobial susceptibility testing, AMR is a significant problem in sub-Saharan Africa (10). Literature indicates that multidrug resistance is prevalent among gram-negative and gram-positive bacterial pathogens causing common infections irrespective of whether community- or hospital-acquired (10, 11). The One Health concept that focusses on consequences, responses, and actions at the animal-human-ecosystems interfaces, is needed to mitigate this issue and hence deserves full support from policymakers and stakeholders. Therefore, the current study aimed to evaluate the antibiotic resistant patterns of pathogens isolated from routine laboratory specimens from 2019 to 2021 at Livingstone Central Hospital (LCH) as a means of understanding the AMR trends at the hospital.

Generally, our study revealed that blood specimens were the most frequently processed specimen with 66.2% followed by urine (33%) and sputum (0.8%) indicating bacteraemia and urinary tract infections as the commonly managed infections at LCH. A similar study conducted in Ndola, Zambia found urine and blood specimens as the common processed specimens at Ndola Teaching Hospital (12). Furthermore, many specimens came from out-patient department (64.7%), and we further noted that the influx of specimens to the laboratory reduced from 35% (for 2019 and 2020) to 30% (for 2021), conceivably because of the COVID19 pandemic that caused Government to put the country on periodic lockdown, thereby reducing the number of patients that accessed health services.

*Staphylococcus aureus* (28.4%) was the commonest isolate followed by *Escherichia coli* (18.2%), *Enterobacter agglomerans* (15.2%), *Klebsiella pneumoniae* (8.6%), and the least isolated bacteria were *Hafnia alvei, Proteus vulgaris*, and *Yersinia enterocolytica* (0.6% apiece). This finding confirmed previous studies that had shown the rising cases of *Staphylococcus aureus* infections in humans and animals in Zambia (12-15). In a quest to understand the susceptibility patterns, the isolated organisms showed high levels of resistance to commonly utilised antibiotics. For instance, Ampicillin (93%) was the least effective drug followed by oxacillin (88%), penicillin (85.6%), co-trimoxazole (81.5%), erythromycin (71.9%), nalidixic acid (68%), ceftazidime (60%), tetracycline (55.1%), and ciprofloxacin (45.9%) whereas the most effective antibiotics were imipenem (14.5%), piperacillin/tazobactam (16.7%) and clindamycin (34.5%). The observed resistance pattern is alarming because most isolates could survive in different classes of antibiotics like penicillins and cephalosporins, macrolides, quinolones, fluoroquinolones, tetracyclines, and sulfonamides, leaving few therapeutic options like carbapenem and lincomycin. Similarly, imipenem was reported to be the most effective drug on uropathogens in India (16), and on *Staphylococcus aureus* and *Staphylococcus pseudintermedius* isolates from the veterinary hospitals’ in- and outpatients and the environment in Zambia (13).

Several studies reveal that sepsis due to bacterial infections emanates from urinary tract infections which are more prevalent in women (17-19). So, we expected to observe the similar trend in this study. Surprisingly, the potency of nitrofurantoin on *Enterobacter agglomerans* (p=0.041), cotrimoxazole on CoNS (p=0.044), cefotaxime on *Klebsiella oxytoca* (p=0.026), and cefuroxime on *Escherichia coli* (p=0.011) were significantly reduced in male patients. This observation required further characterization of blood and urine isolates to clearly understand this phenomenon.

Furthermore, the potency of chloramphenicol on *Enterobacter agglomerans* (p=0.002) and ciprofloxacin on *Klebsiella pneumoniae* (p=0.021) were significantly high among in-patients, which agrees with another study done in Kuwait (20). Finally, cotrimoxazole potency on *Staphylococcus aureus* isolates from blood specimens were significantly (p=0.005) low. Therefore, the use of antibiotics especially from male admitted patients should always be proven by susceptibility testing. The resistant trend observed in this study clearly indicate possible prevalence of extended spectrum beta lactamases (ESBLs) (21), and *Enterobacter agglomerans, Klebsiella pneumonia* and *Staphylococcus aureus* are clinically important pathogens that can develop multidrug resistance and cause difficult to treat infections in all age groups (22, 23). Imipenem is a carbapenem that shows broad-spectrum activity against gram-positive, gram-negative, and anaerobic bacteria, and is active against cephalosporin-resistant Enterobacteriaceae producing ESBLs, whereas piperacillin/tazobactam is the combination of a fourth generation, extended-spectrum penicillin and a beta-lactamase inhibitor that is also effective against β-lactamase producing penicillin-resistant bacterial species (24, 25).

Due to the wide spread of antibiotic resistance resulting from ESBL producing *Enterobacteriaceae*, carbapenems should normally be reserved as alternative treatment for such infections (26) but our study found that both imipenem and piperacillin/tazobactam were broadly used in almost all infections. Despite imipenem being observed as the most effective antibiotics at this LCH, there could be chances of irrational use of the antibiotics subsequently posing a risk of developing carbapenem-resistant bacterial strains. Also, the resistant pattern of isolated strains suggests prevalence of ESBL-producing Enterobacteriaceae that warrant further investigations. To circumvent the issues of carbapenem resistant Enterobacteriaceae and ESBL-producing Enterobacteriaceae, antimicrobial stewardship program should be implemented to curb purported irrational use of antibiotics and enhance adherence to proposed guidelines on antimicrobial use (27). Additionally, the need for more studies (especially molecular studies) to further assess the prevalence of carbapenemases and redesign the empiric therapy to safeguard few potent carbapenems cannot be over emphasized.

### Characterization of bacterial pathogens from paediatric department

Antimicrobial resistance impacts all population and the increasing trend of drug resistant infections in infants and children is usually unrecognized while some studies have shown prevalence of ESBL producing organisms (28, 29). Because of this scarcity of AMR data in children, we decided the characterize bacteria isolates from paediatric department specimens.

We discovered that most specimens were blood and urine that came from in-patient department (74.5%) and there were little variations in the influx of specimens as 36.3% (2019), 28.4% (2020) and 35.3% (2021) were recorded. The most isolated bacterium was *Staphylococcus aureus* (43.1%) followed by *Escherichia coli* (16.7%), *Enterobacter agglomerans* (15.7%), *Klebsiella pneumoniae* (6.9%), *Klebsiella oxytoca* (3.9%), and *Citrobacter freundii* (2.9%). The study revealed that ampicillin (95.5%) was the least effective drug followed by oxacillin (87.5%), penicillin (84.8%), co-trimoxazole (82.5%), erythromycin (74.2%), and nalidixic acid (60%) whereas the most effective antibiotics were imipenem (7.7%), nitrofurantoin (29.6%) and cefotaxime (34.1%). An increasing prevalence of uropathogen resistance to ceftriaxone, cefuroxime, amoxicillin/clavulanate, and ampicillin had been reported in children (30). Therefore, prescribing amoxiclav, cotrimoxazole and nalidixic acid in children should be done cautiously with susceptibility testing.

### Characterization of bacterial isolates from in-patient department

Hospital acquired infections (HAIs) are of major safety concern for both health care providers and the patients (31). These infections are usually acquired after hospitalization and manifest 48-72 hours after admission to the hospital. Even though it was difficult to determine whether the infections were hospital-acquired or not, due to the retrospective nature of our study, we thought of having a glimpse on the resistant levels of bacteria isolated from IPD. We noticed that more blood and urine specimens came from female patients. The most isolated bacterium was *Staphylococcus aureus* (28.2%), followed by *Escherichia coli* (18.1%), *Enterobacter agglomerans* (14.7%), *Klebsiella pneumoniae* (6.8%), CoNS (4.5%), and *Klebsiella oxytoca* (4.5%). The study revealed that ampicillin (93.4%) was the least effective drug followed by penicillin (83%), co-trimoxazole (82.5%), oxacillin (80%), erythromycin (74.4%), nalidixic acid (70.7%), cefuroxime (60%), and tetracycline (48.5%) whereas the most effective antibiotics were norfloxacin (18.2%), imipenem (18.5%) and clindamycin (28.6%). The picture of multidrug resistant strains was observed amongst IPD specimens and this calls for an in-depth analysis of antibiotic susceptibility patterns of nosocomial pathogens. The prevalence of gram-negative HAI- and carbapenem resistant-pathogens have been reported across the globe (32, 33) and the cases are steadily increasing, posing a risk on the cost and effective management of HAIs. Therefore, antimicrobial stewardship programs (27, 31) should be implemented in referral hospitals in Zambia and beyond.

In conclusion, our study has revealed the importance of periodic monitoring of antibiotic resistance patterns in hospitals. The study discovered *Staphylococcus aureus, Escherichia coli, Enterobacter agglomerans* and *Klebsiella pneumoniae* as frequently isolated bacteria from blood and urine specimens with highly resistance to ampicillin, penicillin, oxacillin, co-trimoxazole, and erythromycin but imipenem was the most effective antibiotic. Similarly, paediatric and IPD isolated strains presented with multidrug resistance to commonly used antibiotics at LCH. Therefore, more studies to establish the prevalence of ESBL- and carbapenemase-producing bacteria for empiric therapy redesigning are warranted as monitoring data on antimicrobial susceptibility of common bacterial organisms is crucial for decision-making and quick detection of AMR at hospital levels.

## Data Availability

data adequately presented

## Acknowledgement

We would like to sincerely thank all members of staff in the Laboratory department at Livingstone Central Hospital for their work in generating data used in this study.

## References

1. Mancuso G, Midiri A, Gerace E, Biondo C. Bacterial Antibiotic Resistance: The Most Critical Pathogens. Pathogens (Basel, Switzerland). 2021;10(10).

2. Mhondoro M, Ndlovu N, Bangure D, Juru T, Gombe NT, Shambira G, et al. Trends in antimicrobial resistance of bacterial pathogens in Harare, Zimbabwe, 2012-2017: a secondary dataset analysis. BMC Infect Dis. 2019;19(1):746.

3. Bradford PA. Extended-spectrum beta-lactamases in the 21st century: characterization, epidemiology, and detection of this important resistance threat. Clinical microbiology reviews. 2001;14(4):933–51.

4. Bradford PA, Sanders CC. Development of test panel of beta-lactamases expressed in a common Escherichia coli host background for evaluation of new beta-lactam antibiotics. Antimicrobial agents and chemotherapy. 1995;39(2):308–13.

5. Saravanan M, Ramachandran B, Barabadi H. The prevalence and drug resistance pattern of extended spectrum β–lactamases (ESBLs) producing Enterobacteriaceae in Africa. Microbial Pathogenesis. 2018;114:180–92.

6. Tracking CRE in the United States [Internet]. Centers for Disease Control and Prevention. 2019 [cited 14 July 2020]. Available from: https://www.cdc.gov/hai/organisms/cre/trackingcre.html.

7. New Antibiotics Needed to Fight Nightmare Superbug [Internet]. https://www.pewtrusts.org/en/research-and-analysis/data-visualizations/2020/new-antibiotics-needed-to-fight-nightmare-superbug. 2020 [cited 14 July 2020]. Available from: https://www.pewtrusts.org/-/media/assets/2020/05/cre_v4.pdf.

8. Bell BG, Schellevis F, Stobberingh E, Goossens H, Pringle M. A systematic review and meta-analysis of the effects of antibiotic consumption on antibiotic resistance. BMC Infect Dis. 2014;14:13.

9. CSLI. Performance Standards for antimicrobial susceptibility testing. 26 ed. Wayne PA: Clinical and laboratory standards institute; 2016 8 April 2022. 252 p.

10. Ntirenganya C, Manzi O, Muvunyi CM, Ogbuagu O. High prevalence of antimicrobial resistance among common bacterial isolates in a tertiary healthcare facility in Rwanda. Am J Trop Med Hyg. 2015;92(4):865–70.

11. Leopold SJ, van Leth F, Tarekegn H, Schultsz C. Antimicrobial drug resistance among clinically relevant bacterial isolates in sub-Saharan Africa: a systematic review. The Journal of antimicrobial chemotherapy. 2014;69(9):2337–53.

12. Chanda W, Manyepa M, Chikwanda E, Daka V, Chileshe J, Tembo M, et al. Evaluation of antibiotic susceptibility patterns of pathogens isolated from routine laboratory specimens at Ndola Teaching Hospital: A retrospective study. PLOS ONE. 2019;14(12):e0226676.

13. Youn JH, Park YH, Hang’ombe B, Sugimoto C. Prevalence and characterization of Staphylococcus aureus and Staphylococcus pseudintermedius isolated from companion animals and environment in the veterinary teaching hospital in Zambia, Africa. Comparative immunology, microbiology and infectious diseases. 2014;37(2):123–30.

14. Mutalange M, Yamba K, Kapesa C, Mtonga F, Banda M, Muma JB, et al. Vancomycin Resistance in Staphylococcus aureus and Enterococcus Species isolated at the University Teaching Hospitals, Lusaka, Zambia: Should We Be Worried? University of Zambia Journal of Agricultural and Biomedical Sciences. 2021;5(1).

15. Mulongo T, Kamvuma K, Phiri CN, Mulemena JA, Chanda W. Elevators and staircase handrails as potential sources of nosocomial pathogens at Ndola Teaching Hospital, Zambia. 2021.

16. Somashekara SC, Deepalaxmi S, Jagannath N, Ramesh B, Laveesh MR, Govindadas D. Retrospective analysis of antibiotic resistance pattern to urinary pathogens in a Tertiary Care Hospital in South India. Journal of basic and clinical pharmacy. 2014;5(4):105.

17. Wagenlehner FME, Pilatz A, Weidner W, Naber KG. Urosepsis: Overview of the Diagnostic and Treatment Challenges. Microbiol Spectr. 2015;3(5).

18. Nicolle LE. Urinary tract infection. Crit Care Clin. 2013;29(3):699–715.

19. Petrosillo N, Granata G, Boyle B, Doyle MM, Pinchera B, Taglietti F. Preventing sepsis development in complicated urinary tract infections. Expert Rev Anti Infect Ther. 2020;18(1):47–61.

20. Alali WQ, AlFouzan W, Dhar R. Prevalence of antimicrobial resistance in Gram-negative clinical isolates from a major secondary hospital in Kuwait: a retrospective descriptive study. Germs. 2021;11(4):498–511.

21. Agga GE, Silva PJ, Martin RS. Detection of Extended-Spectrum Beta-Lactamase-Producing and Carbapenem-Resistant Bacteria from Mink Feces and Feed in the United States. Foodborne pathogens and disease. 2021;18(7):497–505.

22. Büyükcam A, Tuncer Ö, Gür D, Sancak B, Ceyhan M, Cengiz AB, et al. Clinical and microbiological characteristics of Pantoea agglomerans infection in children. Journal of Infection and Public Health. 2018;11(3):304–9.

23. Luo K, Tang J, Qu Y, Yang X, Zhang L, Chen Z, et al. Nosocomial infection by Klebsiella pneumoniae among neonates: a molecular epidemiological study. The Journal of hospital infection. 2021;108:174–80.

24. LiverTox. Piperacillin-Tazobactam. LiverTox: Clinical and Research Information on Drug-Induced Liver Injury. Bethesda (MD): National Institute of Diabetes and Digestive and Kidney Diseases; 2012.

25. Doi Y. Ertapenem, Imipenem, Meropenem, Doripenem, and Aztreonam. Mandell, Douglas, and Bennett’s Principles and Practice of Infectious Diseases 9th ed Elseviver. 2020:285–90.

26. Mitgang EA, Hartley DM, Malchione MD, Koch M, Goodman JL. Review and mapping of carbapenem-resistant Enterobacteriaceae in Africa: Using diverse data to inform surveillance gaps. International Journal of Antimicrobial Agents. 2018;52(3):372–84.

27. MacDougall C, Polk RE. Antimicrobial stewardship programs in health care systems. Clinical microbiology reviews. 2005;18(4):638–56.

28. Medernach RL, Logan LK. The Growing Threat of Antibiotic Resistance in Children. Infect Dis Clin North Am. 2018;32(1):1–17.

29. Williams PCM, Isaacs D, Berkley JA. Antimicrobial resistance among children in sub-Saharan Africa. The Lancet Infectious Diseases. 2018;18(2):e33–e44.

30. Nadav Graif SA, and Avi Peretz. Trends in Distribution and Antibiotic Resistance of Bacteria Isolated from Urine Cultures of Children in Northern Israel Between 2010 and 2017. Microbial Drug Resistance. 2020;26(11):1342–9.

31. Mehta Y, Gupta A, Todi S, Myatra S, Samaddar DP, Patil V, et al. Guidelines for prevention of hospital acquired infections. Indian J Crit Care Med. 2014;18(3):149–63.

32. Ogwang M, Paramatti D, Molteni T, Ochola E, Okello TR, Ortiz Salgado JC, et al. Prevalence of hospital-associated infections can be decreased effectively in developing countries. Journal of Hospital Infection. 2013;84(2):138–42.

33. Phu VD, Wertheim HFL, Larsson M, Nadjm B, Dinh Q-D, Nilsson LE, et al. Burden of Hospital Acquired Infections and Antimicrobial Use in Vietnamese Adult Intensive Care Units. PLOS ONE. 2016;11(1):e0147544.

